# LIFETIME JEM-DERIVED OCCUPATIONAL EXPOSURE BURDEN AND ODDS OF PARKINSON’S DISEASE AND PARKINSONISM IN A MATCHED CASE-CONTROL STUDY

**DOI:** 10.64898/2026.03.16.26348171

**Authors:** Freeman Lewis, Stefano Renzetti, Natalie Goulett, Somaiyeh Azmoun, Vaishnavi Sundar, Mahnoor Ali, Luciana Pitta, Daniel Shoieb, Margherita Caci, Stefano Borghesi, Loredana Covolo, Manuela Oppini, Umberto Gelatti, Alessandro Padovani, Andrea Pilotto, Fulvio Pepe, Marinella Turla, Patrizia Crippa, Paolo Boffetta, Luca Pani, Roel Vermeulen, Hans Kromhout, Luca Lambertini, Elena Colicino, Donatella Placidi, Roberto Lucchini

## Abstract

**Background/Objectives:** Occupational exposure to neurotoxicants, including pesticides and metals, has been implicated in Parkinson’s disease (PD) and Parkinsonism. However, the cumulative impact of lifetime occupational exposures remains insufficiently characterized. This study evaluated whether semi-quantitative, ALOHA+ Job-Exposure Matrix (ALOHA+-JEM) – derived cumulative occupational exposures were associated with PD and Parkinsonism.

**Methods:** We conducted a hospital-based matched case–control study in Brescia, Italy, (n = 668; 334 PD/Parkinsonism cases and 334 matched controls), 1:1 matched on sex, age, and lifetime occupational duration. Lifetime occupational histories were coded using ISCO classification and linked to the ALOHA+-JEM to derive cumulative exposure metrics (unexposed/low/high) for major occupational agent families. Associations were estimated using conditional logistic regression adjusting for smoking, parental history of PD/tremor, and SNCA rs356219 genotype. Weighted Quantile Sum (WQS) regression was applied as a secondary analysis to explore co-exposure structure and relative contributions to a composite occupational exposure burden index.

**Results:** Parental history of PD or tremor (OR = 4.55, 95% CI: 2.44–8.48; q < 0.001) and the SNCA rs356219 CC genotype (OR = 2.17, 95% CI: 1.33–3.52; q = 0.013) were associated with disease, consistent with established risk factors. High cumulative pesticide exposure was nominally associated with increased odds of combined PD and Parkinsonism (OR = 2.98, 95% CI: 1.23–7.25) and PD alone (OR = 3.56, 95% CI: 1.25–10.15); however, these associations did not remain statistically significant after false discovery rate correction. Other exposure families were not consistently associated with disease. In WQS analyses, the composite occupational exposure burden index showed modest positive association with combined PD and Parkinsonism (OR = 1.15, 95% CI: 1.00–1.30). All pesticides and metals contributed most strongly to the index, although estimates were imprecise, particularly in the subgroup analysis.

**Conclusions:** ALOHA+-JEM-derived cumulative occupational exposures showed suggestive but modest associations with PD and Parkinsonism, with pesticides contributing most consistently across analyses. These findings should be interpreted cautiously given the semi-quantitative nature of exposure assessment and limited statistical significance after multiple testing correction. Overall, the results support a potential role for cumulative occupational exposures in Parkinsonian disorders, while highlighting the need for studies incorporating more resolved exposure assessment approaches.

## 1. Introduction

Humans spend a substantial proportion of their lives in indoor environments, including both residential and occupational settings. A considerable amount of research has focused on residential exposures; however, occupational environments remain an important and comparatively less studied source of potentially hazardous exposures (Klepeis et al., 2001; Liu et al., 2023). National time-use surveys indicate that U.S. adults spend roughly 70% of their waking hours, nearly 16 hours per day, at home, contributing to this emphasis on residential environments (Sharkey, 2024). In contrast, occupational exposures remain substantial: approximately 80% of full-time employees spend an average of 8.4 hours per weekday at work and the expected duration of working life has increased globally (Eurostat, 2025; U.S. Bureau of Labor Statistics, 2025). Although remote and hybrid work have reduced workplace exposure for some groups, these trends are occupation-specific and are less common in industrial, agricultural, manufacturing, and service occupations. Consequently, many workers, particularly in manual and technical occupations, continue to experience prolonged and potentially hazardous occupational exposures, emphasizing the importance of accounting for workplace exposures in cumulative exposure assessment.

While residential addresses and geographical information systems (GIS) are commonly leveraged to operationalize environmental exposure assessment, occupational environments, particularly in industries such as manufacturing and welding, present complex exposure scenarios that are not readily captured by residential proxies (Hoek et al., 2024; Nuckols John et al., 2004). In these settings, workers may experience substantially higher exposure intensity, longer duration, and repeated contact across the working life when compared to environmental exposures. Consequently, occupational exposures may account for a disproportionately large share of an individual’s overall internal exposure burden (Azmoun et al., 2022; Azmoun et al.,2025; Barbey et al., 2024; Peters et al., 2024; Quintero Santofimio et al., 2024). Yet despite the potential magnitude and public health relevance, quantifying work exposures at the population level remains challenging due to a lack of job-specific measurement data and variability in exposures over time across working life. To address this gap, general population Job-Exposure Matrices (JEMs) offer a standardized, validated framework that links occupational titles, classified by international coding systems (i.e., International Standard Classification of Occupations 1968 or 1988 [ISCO-68 and ISCO-88]), to probability and intensity of occupational exposure in the general population (Choi, 2020; Descatha et al., 2022; Fevotte et al., 2011; Mannetje and Kromhout, 2003; Peters et al., 2022). By cross-referencing occupational titles with probabilistic or semi-quantitative exposure estimates, JEMs enable population-level inference of chemical exposures in a cost-efficient and systematic manner.

In the present study, we applied the ALOHA+-JEM to derive semi-quantitative estimates of occupational exposures to known neurotoxicants and assess their associations with Parkinson’s disease (PD) and Parkinsonism within an occupational epidemiology framework (van der Market al., 2014). PD is a progressive neurodegenerative disorder primarily affecting dopaminergic neurons of the substantia nigra, whereas Parkinsonism encompasses a broader spectrum of motor syndromes that share overlapping clinical features but may arise from diverse etiologies, including toxicant exposure (Hayes, 2019). Epidemiologic and toxicologic evidence has previously proven pesticides, notably organochlorines and paraquat, in PD pathogenesis, while metals such as manganese (Mn), lead (Pb), and copper (Cu) are more consistently associated with Parkinsonism (Stoccoro and Coppedè, 2024). Additional workplace agents, such as solvents (i.e., trichloroethylene), have also been linked to increased risk of PD, whereas gases and fumes and mineral dust remain incompletely characterized but are considered to have neurotoxic potential (Dorsey and Bloem, 2024; Goldman et al., 2023). These agents were selected because they represent modifiable, non-genetic risk factors relevant to occupational settings.

Although the neurotoxic profile of many of these agents is well established, their cumulative effects across the working life remain poorly quantified. Based on consistent epidemiological and experimental evidence, we hypothesized that higher cumulative occupational exposure to pesticides and metals would be associated with increased risk of Parkinson’s disease (PD) and Parkinsonism (Liu et al., 2025; Lucchini and Tieu, 2023). To evaluate this hypothesis, we applied the ALOHA+-JEM to lifetime occupational histories in a hospital-based case–control study. In addition, we explored associations for other occupational exposure families—including aromatic and chlorinated solvents, gases and fumes, and mineral dust—as secondary, exploratory analyses, given less consistent prior evidence. This approach enables evaluation of primary a priori hypotheses while providing an exploratory assessment to capture cumulative exposure relevant to the long latency of disease development (Lewis et al., 2025; Lucchini et al., 2007; Lucchini et al., 2020).

## 2. Methods

### 2.1. Study population

The study population has been described in detail previously; in brief, a hospital-based case–control study was conducted in the Province of Brescia, Northern Italy (Lucchini et al., 2020). This study population represents a unique regional cohort residing in an area with long-standing metallurgical activity, including ferromanganese production dating back to the early 1900s. This historically industrialized area has also developed intensive agricultural activity over the same period, accompanied by increasing pesticide use across a variety of crops. Participant recruitment occurred across four movement-disorder centers—ASST-Spedali Civili of Brescia, Esine Hospital of Valcamonica, Poliambulanza Foundation, and Ancelle Domus Salutis—under a common diagnostic protocol and centralized ethical oversight. Participants were consecutively enrolled during 2012-2015 to minimize selection bias and ensure comparability across centers. Participants were not followed longitudinally after enrollment. The patient sample size included 430 patients with Parkinson’s disease (PD) or Parkinsonism (38% females) and 446 hospital-based controls without diagnosable neurological disease. Summary demographic and clinical characteristics are presented in Table 1. For the present analysis, 668 subjects (334 PD/Parkinsonism cases and 334 matched controls) with complete occupational and covariate data were included. The diagnostic classification extended beyond idiopathic PD to the broader category of Parkinsonism, defined by the presence of at least two of the following cardinal motor features: bradykinesia/akinesia, rigidity, tremor, or postural instability, while excluding cases of vascular, iatrogenic, or traumatic origin. Diagnoses were made by movement-disorder neurologists at each site (authors AP, MCR, AP at Spedali Civili, FP at Poliambulanza Foundation, MT at Esine Hospital, and PC at Ancelle Brescia) according to standardized clinical criteria and reviewed by a single senior neurologist to ensure diagnostic consistency. Non-Parkinsonian controls were recruited from the Dermatology, Orthopedics, Ophthalmology, and Otorhinolaryngology clinics within the same hospitals to ensure comparable catchment areas and residential distributions. Controls were matched to cases on sex, age (±5 years), and occupational life up to the case’s diagnosis date, with one control per case (1:1 matching ratio). For each matched pair, the control’s occupational follow-up was truncated at the corresponding case diagnosis date. While selection bias cannot be entirely excluded, matching occupational duration helped mitigate differences in exposure opportunity between cases and controls. Trained interviewers administered structured questionnaires capturing lifetime occupational histories (job titles, ISCO codes, industry, tasks, start and end dates, and duration), demographics, smoking status, and family history of PD (parents only, to minimize recall bias). Questionnaires were novel and not adapted from previously validated instruments. The same questionnaire was administered to both cases and controls using standardized protocols to ensure comparability of data collection. Blood samples were obtained from all participants at enrollment for SNCA (rs356219) genotyping (details in Section 2.3 Covariates). Structured questionnaires and blood sampling were consistent with the period for which patients were enrolled. The study was conducted in accordance with the Declaration of Helsinki and approved by the Ethics Committee of the Civil Hospital in Brescia, Italy (PARKGENEAMB protocol, approval granted 4 September 2012), and by the Institutional Review Board of Florida International University (protocol code IRB-22-0246, approval granted 27 May 2022). Written informed consent was obtained from all participants prior to enrollment. All primary data can be found in Supplementary Table S1.

**Table 1.** Demographic, clinical, and genetic characteristics of study participants by diagnostic group (controls, Parkinson’s disease [PD], and Parkinsonism)

|  | Controls<br>(N=334) | PD<br>(N=260) | Parkinsonism<br>(N=74) | Total<br>(N=668) |
| --- | --- | --- | --- | --- |
| Age |  |  |  |  |
| Mean (SD) | 69.1 (9.8) | 70.5 (9.8) | 74.1 (7.5) | 70.2 (9.7) |
| Sex |  |  |  |  |
| Female, n (%) | 119 (35.6%) | 93 (35.8%) | 26 (35.1%) | 238 (35.6%) |
| Male, n (%) | 215 (64.4%) | 167 (64.2%) | 48 (64.9%) | 430 (64.4%) |
| Occupational Duration (Years) |  |  |  |  |
| Mean (SD) | 34 (13) | 33.4 (13.1) | 32.1 (14.7) | 33.7 (13.3) |
| Family History of PD or Tremor |  |  |  |  |
| Yes, n (%) | 15 (4.5%) | 45 (17.2%) | 12 (16.7%) | 72 (10.8%) |
| No, n (%) | 319 (95.5%) | 217 (82.8%) | 60 (83.3%) | 596 (89.2%) |
| SNCA rs356219<sup>b</sup> |  |  |  |  |
| TT, n (%) | 145 (43.4%) | 91 (34.7%) | 23 (31.9%) | 259 (38.8%) |
| TC, n (%) | 147 (44.0%) | 122 (46.6%) | 34 (47.2%) | 303 (45.4%) |
| CC, n (%) | 42 (12.6%) | 49 (18.7%) | 15 (20.8%) | 106 (15.9%) |
| Ever Smoker |  |  |  |  |
| Ever, n (%) | 161 (48.2%) | 103 (39.6%) | 34 (45.9%) | 298 (44.6%) |
| Never, n (%) | 173 (51.8%) | 157 (60.4%) | 40 (54.1%) | 370 (55.4%) |
| Alcohol Consumption |  |  |  |  |
| Ever, n (%) | 232 (69.5%) | 177 (67.6%) | 54 (75.0%) | 463 (69.3%) |
| Never, n (%) | 102 (30.5%) | 85 (32.4%) | 18 (25.0%) | 205 (30.7%) |
| Born in Brescia Province |  |  |  |  |
| Yes, n (%) | 284 (85.5%) | 242 (92.7%) | 67 (94.4%) | 593 (89.3%) |
| No, n (%) | 48 (14.5%) | 19 (7.3%) | 4 (5.6%) | 71 (10.7%) |
| Comorbidities |  |  |  |  |
| Diabetes |  |  |  |  |
| Yes, n (%) | 66 (19.9%) | 25 (9.6%) | 4 (5.6%) | 95 (14.3%) |
| No, n (%) | 265 (80.1%) | 235 (90.4%) | 68 (94.4%) | 568 (85.7%) |
| Stroke |  |  |  |  |
| Yes, n (%) | 4 (1.2%) | 12 (4.6%) | 2 (2.8%) | 18 (2.7%) |
| No, n (%) | 327 (98.8%) | 248 (95.4%) | 70 (97.2%) | 645 (97.3%) |
| Hypertension |  |  |  |  |
| Yes, n (%) | 197 (59.5%) | 139 (53.3%) | 34 (47.2%) | 370 (55.7%) |
| No, n (%) | 134 (40.5%) | 122 (46.7%) | 38 (52.8%) | 294 (44.3%) |
| Leukemia |  |  |  |  |
| Yes, n (%) | 2 (0.6%) | 0 (0%) | 0 (0%) | 2 (0.3%) |
| No, n (%) | 329 (99.4%) | 260 (100.0%) | 72 (100.0%) | 661 (99.7%) |
| Heart Disease |  |  |  |  |
| Yes, n (%) | 88 (26.5%) | 62 (23.8%) | 22 (30.6%) | 172 (25.9%) |
| No, n (%) | 244 (73.5%) | 198 (76.2%) | 50 (69.4%) | 492 (74.1%) |
| Liver Disease |  |  |  |  |
| Yes, n (%) | 18 (5.4%) | 15 (5.8%) | 3 (4.2%) | 36 (5.4%) |
| No, n (%) | 313 (94.6%) | 245 (94.2%) | 69 (95.8%) | 627 (94.6%) |
| Kidney Disease |  |  |  |  |
| Yes, n (%) | 22 (6.6%) | 12 (4.6%) | 4 (5.6%) | 38 (5.7%) |
| No, n (%) | 309 (93.4%) | 248 (95.4%) | 68 (94.4%) | 625 (94.3%) |
| Thyroid Disease |  |  |  |  |
| Yes, n (%) | 24 (7.3%) | 16 (6.2%) | 9 (12.5%) | 49 (7.4%) |
| No, n (%) | 306 (92.7%) | 244 (93.8%) | 63 (87.5%) | 613 (92.6%) |

Continuous variables are presented as mean ± standard deviation (SD), and categorical variables as counts and percentages. Age and sex were matching variables in the study design. SNCA rs356219 genotype coded as TT (reference), TC (heterozygous), and CC (homozygous variant).

### 2.2. Semi-quantitative ALOHA+-JEM (Job Exposure Matrix)

Job histories were collected by structured interviews for every job held, recording the job title, main tasks, employer or industry, and start and end years. All jobs were coded to four-digit International Standard Classification of Occupations 2008, unit groups by trained coders using the International Labour Organization (ILO) guidelines for structure and coding, with emphasis on task and duty information to resolve ambiguous titles (International Labour Organization, 2012). The ISCO-08 codes were then recoded to the International Standard Classification of Occupations, 1988 (ISCO-88) framework through an official ILO crosswalk to enable linkage with the ALOHA+ Job-Exposure Matrix (ALOHA+-JEM) (Humlum, 2021). Coding was performed independently of case–control status, and discrepancies were resolved by consensus review. For descriptive analyses of occupational class, participants’ job titles were categorized according to the major occupational groups including Managers, Professionals, Technicians and Associate Professionals, Clerical Support Workers, Services and Sales Workers, Craft and Related Trades Workers, Plant and Machine Operators and Assemblers, and Elementary Occupations (International Labour Organization, 2012). For each coded job, the ALOHA+-JEM (developed by Utrecht University, Netherlands) assigned semi-quantitative exposure levels (unexposed, low, or high) across agent families including mineral and inorganic dusts, aromatic and chlorinated solvents, other organic solvents, pesticides and metals (van der Mark et al., 2014). Job duration in years was calculated using job start and end dates, with end dates truncated at the year of diagnoses for cases and the corresponding matched year for controls. Cumulative occupational exposure was quantified using job-level intensity ratings from the ALOHA+-JEM (unexposed = 0, low = 1, high = 4). For each exposure agent, the intensity score assigned to each job was multiplied by the number of years the participant held that job. These job-specific products were then summed across all jobs in the participant’s occupational history to obtain a cumulative exposure score for each agent. Participants were subsequently classified into three ordinal exposure categories: unexposed (cumulative score = 0), low cumulative exposure (cumulative score > 0 and ≤ the median among exposed participants), and high cumulative exposure (cumulative score > the median among exposed participants). The ALOHA+-JEM assigns exposures at the level of agent families rather than individual chemicals. For each job, a single semi-quantitative exposure level (unexposed, low, or high) is assigned per agent family based on the combination of exposure probability and intensity. Consequently, multiple agents within the same family are not quantified separately or summed; instead, the assigned exposure level reflects an overall classification for that agent group within a given job. Cumulative exposure scores therefore represent the aggregation of job-level exposure estimates across the working life rather than the summation of multiple agents within a single job. The ALOHA+-JEM has been applied in several European epidemiological studies and is designed for developed industrialized settings comparable to Northern Italy (Damerau et al., 2025; Faruque et al., 2020; Peters et al., 2022; van der Mark et al., 2014). In our application, an exposure assignment was conducted independently of disease status using the ALOHA+-JEM version dated 16 November 2020 to ensure reproducibility. All primary data can be found in Supplementary Table S1.

### 2.3. Covariates

Candidate covariates were selected a priori based on established or hypothesized risk factors for PD and Parkinsonism and to control for potential confounding in the association between cumulative occupational exposures and disease status. Covariates included smoking status (ever vs. never), positive family history of PD or tremor (yes vs. no), and SNCA rs356219 genotype (TT, TC, CC), which was previously associated with susceptibility to PD in a previous study conducted on this cohort (Caramiello and Pirota, 2024; Li et al., 2025; Lucchini et al., 2020). These variables were incorporated as adjustment terms in all conditional logistic and weighted quantile sum regression models. For genotyping, DNA was extracted from 0.2 mL of peripheral whole blood using the QIAamp DNA Blood Mini Kit (Qiagen, Hilden, Germany). The SNCA rs356219 polymorphism was genotyped using TaqMan real-time PCR assays (Life Technologies, Carlsbad, CA, USA) following manufacturer protocols. The TaqMan assay for SNCA rs356219 reports T/C alleles on the reverse strand; for comparison with reference databases (Ensembl/1000 Genomes, forward strand A/G), we applied the complement mapping (T→A, C→G) so that TT/TC/CC correspond to AA/AG/GG. For quality control, more than 5% of samples were reanalyzed in independent runs, yielding 100% concordance between duplicates. Genotype frequencies for SNCA rs356219 were concordant with the Tuscans in Italy (TSI) reference (MAF ≈ 0.42) and with the broader European (EUR) super-population (MAF ≈ 0.39) from the 1000 Genomes Project; patterns are in line with prior data from the same cohort (Lucchini et al., 2020). Genotype distributions conformed to Hardy–Weinberg equilibrium (χ² test, p > 0.05), and no significant sex-specific differences in genotype frequencies were observed (χ² test, p > 0.05). The SNCA rs356219 variant was modeled under a co-dominant framework, with the TT genotype set as the reference category and TC and CC representing heterozygous and homozygous variant classes, respectively. All primary data can be found in Supplementary Table S1

### 2.4. Statistical analysis

#### 2.4.1. Descriptive statistics

Descriptive analyses summarized demographic, clinical, genetic, and occupational characteristics of study participants across three diagnostic groups: controls, Parkinson’s disease (PD), and Parkinsonism. Continuous variables (e.g., age) are presented as mean ± standard deviation (SD), and categorical variables—including smoking status (ever/never), family history of PD or tremor (yes/no), SNCA rs356219 genotype categories (TT/TC/CC), comorbid conditions (yes/no), alcohol consumption (ever/never), and birthplace within the Brescia province (yes/no)—are presented as counts and percentages. Occupational cumulative exposure categories (unexposed /low/high) for each agent family derived from the ALOHA+-JEM are similarly summarized as counts and percentages across diagnostic groups. Group comparisons were evaluated using one-way analysis of variance (ANOVA) for continuous variables and chi-squared (χ²) tests for categorical variables; however, corresponding p-values are not presented in the main tables, as these analyses are descriptive and not part of the primary inferential framework.

#### 2.4.2. Primary inferential framework: multivariate conditional logistic regression

Primary analyses evaluated the association between cumulative occupational exposures to agent families (pesticides, metals, aromatic/chlorinated solvents, mineral dust, and gases and fumes) and the risk of Parkinson’s disease (PD) and Parkinsonism using multivariate conditional logistic regression. All occupational exposure families were included simultaneously in the models to account for co-exposure, such that estimates for each exposure represent associations adjusted for other agent families. Models were implemented using the *clogit* function from the *survival* R package with strata (case-control pairs) to account for the 1:1 matched design (matching on age, sex, and occupational duration), yielding odds ratios (ORs) and 95% confidence intervals (CIs) for three prespecified contrasts: (A) PD+Parkinsonism vs Controls (N = 334 controls, 334 cases), (B) PD vs Controls (N = 260 controls, 260 PD), and (C) Parkinsonism vs Controls (N = 74 controls, 74 Parkinsonism). Models were adjusted for smoking status, parental history of PD/tremor, and SNCA rs356219 genotype (CC and TC vs TT); regression p-values were obtained using two-sided Wald z-tests, and where indicated, multiplicity was controlled using the Benjamini–Hochberg false discovery rate (FDR; q < 0.05). All analyses and visualizations were performed in R 4.4.3 (R Core Team, 2025) and Python 3.11.14 within JupyterLab 4.4.10 using the following key packages: survival 3.8.3, dplyr 1.1.4, pandas 2.3.3, numpy 2.3.4, scikitand Mito 0.2.47.

#### 2.4.3. Secondary exploratory analyses: co-exposure structure and WQS regression

To evaluate co-exposure structure, Spearman pairwise correlations (ρ) among working life cumulative exposure to agent families were computed, producing an overall correlation matrix visualized as a heatmap with color intensity representing the strength and direction of correlation (scale −1 to 1). Multi-agent occupational exposure burden indexes were computed using single index weighted quantile sum (WQS) regression implemented via the gWQS package (version 3.0.4), using 100 bootstraps and 100 repeated holdouts (validation fraction = 0.9 per repeat).

Logistic regression models were used in the validation step and included smoking status, parental history of Parkinson’s disease or tremor, and SNCA genotype as covariates. The WQS index was constrained to a positive direction, and component weights were averaged across bootstrap samples and interpreted relative to an equal-weight benchmark. Component weights were estimated using bootstrap sampling in the training step, where regression-based optimization identified the relative contribution of each exposure to the weighted index under a positivity constraint. Because cases and controls were 1:1 matched on age, sex, and occupational duration, each matched set was identified by a unique matching ID. To preserve this matched design during resampling, repeated holdout (RH) partitions were generated by sampling unique matching IDs so that paired observations remained within the same training or validation subset. All analyses and visualizations were performed in R 4.4.3 (R Core Team, 2025) and Python 3.11.14 within JupyterLab 4.4.10 using the following key packages: gWQS 3.0.4, survival 3.8.3, dplyr 1.1.4, pandas 2.3.3, numpy 2.3.4, and Mito 0.2.47.

## 3. Results

### 3.1. Demographic, clinical, and genetic characteristics by diagnostic group

Table 1 reports the demographic, clinical, and genetic characteristics of study participants by diagnostic group (Supplementary Table S1). Age and sex were matching variables in the study design, with age matched within ±5 years; their distributions were therefore broadly comparable across cases and controls. Participants with PD (71.1 ± 9.8 years) and Parkinsonism (71.9 ± 8.0 years) were slightly older than controls (69.1 ± 9.8 years), reflecting residual variation permitted by the matching criteria. A positive family history of PD or tremor was more common among PD (17.2%) and Parkinsonism (12.7%) cases compared with controls (4.5%). The distribution of SNCA rs356219 genotypes showed a higher frequency of the CC genotype among PD (18.7%) and Parkinsonism (20.8%) compared to controls (12.6%). Genotype frequencies were TT = 39%, TC = 45%, and CC = 16%, conformed to Hardy–Weinberg equilibrium (p > 0.05), did not differ by sex (p > 0.05), and were consistent with those previously reported in the same Italian cohort (Lucchini et al., 2020). Alcohol consumption and smoking patterns were similar across diagnostic groups. A higher proportion of PD (92.7%) and parkinsonism (94.4%) cases were born in the Brescia province compared with controls (85.5%). Diabetes and stroke were more prevalent among PD participants, while hypertension was common across all groups without clear differences. Other chronic conditions did not differ meaningfully between groups.

### 3.2. Occupational exposure distribution by diagnostic group derived from ALOHA+-JEM

Table 2 presents participant-level distributions of cumulative occupational exposure categories derived from the ALOHA+-JEM, stratified by diagnostic group and agent family (Supplementary Table S1). Because participants commonly held multiple jobs over their working lifetime, exposure categories reflect cumulative occupational history and were classified as unexposed, low only (i.e., low exposure in one or more jobs and no high exposure), or high (ever) exposure (i.e., high exposure in at least one job). For mineral dust, 48.2% of participants were unexposed, 26.2% had low-only exposure, and 25.6% had high (ever) exposure, with broadly similar distibutions across controls, PD, and Parkinsonism. Comparable distributions were observed for gases and fumes (unexposed = 31.6%, only low = 35.3%, high [ever] = 33.1%). For aromatic/chlorinated solvents, most participants were unexposed (65.1%), while 14.5% had low-only exposure and 20.4% had high (ever) exposure. Metals showed a similar distribution (unexposed = 71.9%, low only = 14.1%, high [ever] = 14.1%). Pesticide exposure was least prevalent overall, with 86.2% unexposed, 7.2% with low-only exposure, and 6.6% with high (ever) exposure.

**Table 2.**
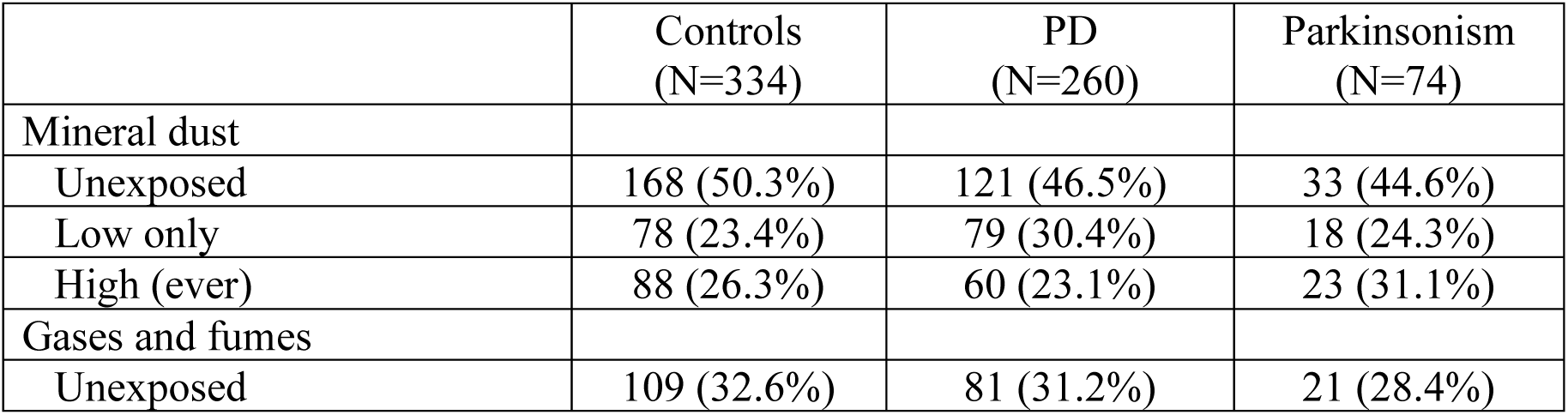
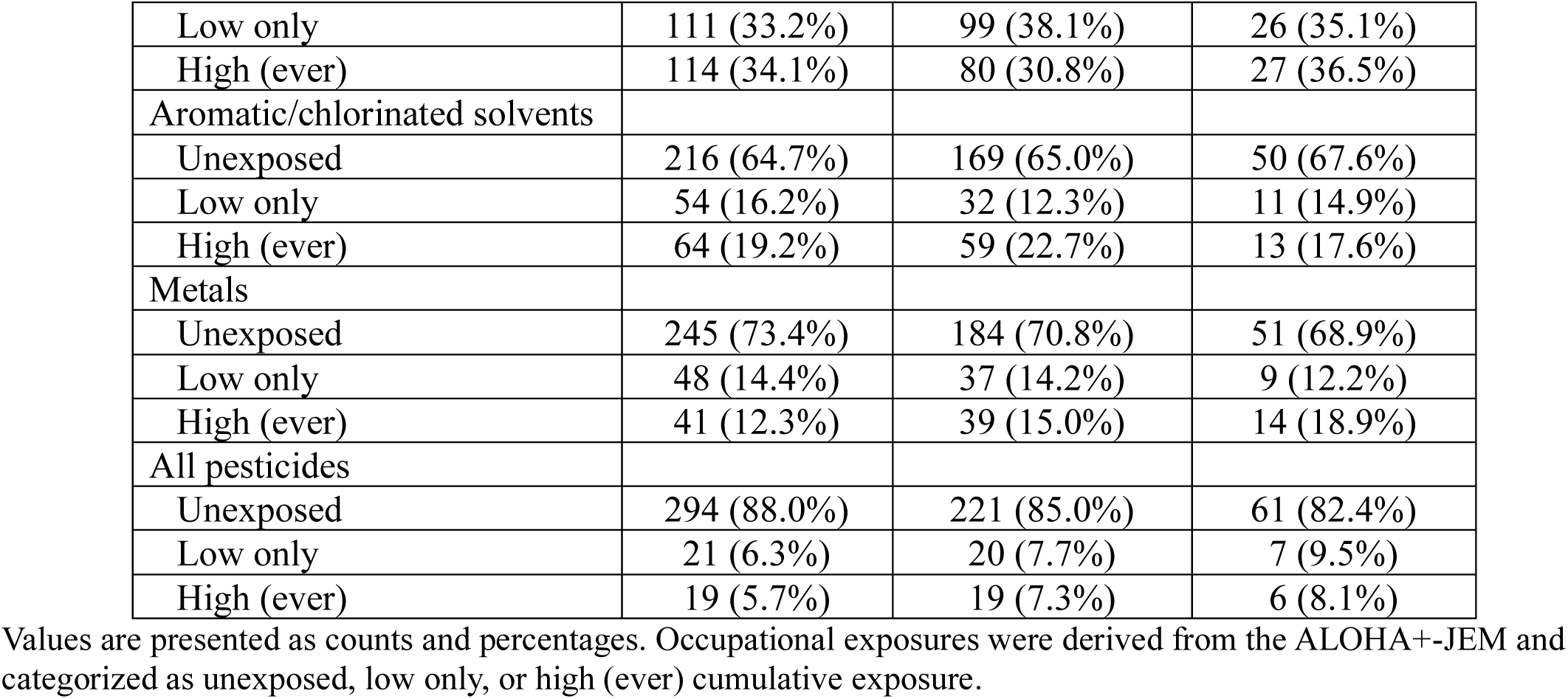
Occupational exposure distributions derived from the ALOHA+-JEM within agent families stratified by diagnostic group (Controls, Parkinson’s disease [PD], and Parkinsonism)

|  | Controls<br>(N=334) | PD<br>(N=260) | Parkinsonism<br>(N=74) |
| --- | --- | --- | --- |
| Mineral dust |  |  |  |
| Unexposed | 168 (50.3%) | 121 (46.5%) | 33 (44.6%) |
| Low only | 78 (23.4%) | 79 (30.4%) | 18 (24.3%) |
| High (ever) | 88 (26.3%) | 60 (23.1%) | 23 (31.1%) |
| Gases and fumes |  |  |  |
| Unexposed | 109 (32.6%) | 81 (31.2%) | 21 (28.4%) |
| Low only | 111 (33.2%) | 99 (38.1%) | 26 (35.1%) |
| High (ever) | 114 (34.1%) | 80 (30.8%) | 27 (36.5%) |
| Aromatic/chlorinated solvents |  |  |  |
| Unexposed | 216 (64.7%) | 169 (65.0%) | 50 (67.6%) |
| Low only | 54 (16.2%) | 32 (12.3%) | 11 (14.9%) |
| High (ever) | 64 (19.2%) | 59 (22.7%) | 13 (17.6%) |
| Metals |  |  |  |
| Unexposed | 245 (73.4%) | 184 (70.8%) | 51 (68.9%) |
| Low only | 48 (14.4%) | 37 (14.2%) | 9 (12.2%) |
| High (ever) | 41 (12.3%) | 39 (15.0%) | 14 (18.9%) |
| All pesticides |  |  |  |
| Unexposed | 294 (88.0%) | 221 (85.0%) | 61 (82.4%) |
| Low only | 21 (6.3%) | 20 (7.7%) | 7 (9.5%) |
| High (ever) | 19 (5.7%) | 19 (7.3%) | 6 (8.1%) |

Values are presented as counts and percentages. Occupational exposures were derived from the ALOHA+-JEM and categorized as unexposed, low only, or high (ever) cumulative exposure.

### 3.3. Correlation structure of cumulative occupational exposures between agent families

Spearman correlation analyses of cumulative occupational exposures demonstrated similar correlation structures across cases (PD and Parkinsonism) and controls, with moderate to strong positive correlations observed among several agent families (Figure 1). Among cases, the strongest correlations were observed between mineral dust and gases and fumes (ρ = 0.78) and between metals and aromatic/chlorinated solvents (ρ = 0.77). Moderate correlations were also observed between metals and mineral dust (ρ = 0.62), metals and gases and fumes (ρ = 0.49), and aromatic/chlorinated solvents and mineral dust (ρ = 0.45). Correlations involving all pesticides were generally weaker, ranging from zero to moderate (ρ = 0.01–0.47). Among controls, the strongest correlations occurred between mineral dust and gases and fumes (ρ = 0.77) and between metals and aromatic/chlorinated solvents (ρ = 0.69). Moderate correlations were also present between metals and mineral dust (ρ = 0.57), metals and gases and fumes (ρ = 0.46), and aromatic/chlorinated solvents and mineral dust (ρ = 0.39). As in cases, all pesticides showed weak to moderate correlations with other agent families (ρ = 0.03–0.43). The results of the Spearman correlation analysis are presented in Supplementary Table S2.

**Figure 1.**
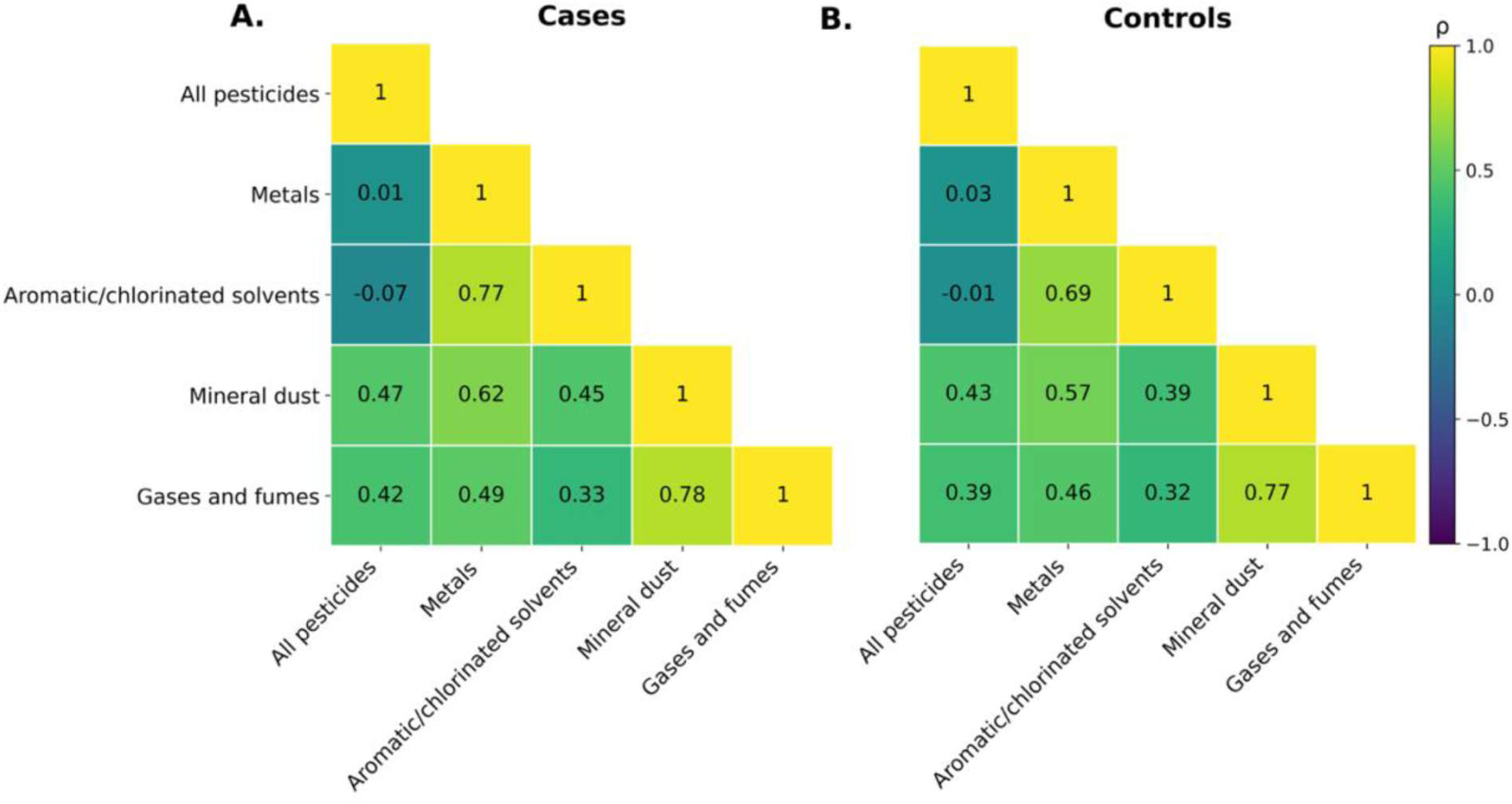
Spearman correlation matrices for cumulative occupational exposures derived from the ALOHA+-JEM. Panels display pairwise Spearman correlation coefficients (ρ) among lifetime cumulative exposure scores to all pesticides, metals, aromatic/chlorinated solvents, mineral dust, and gases and fumes, stratified by cases (A) and controls (B). Cell values indicate the magnitude of ρ for each exposure pair. The color scale represents the strength and direction of correlations, ranging from −1 (negative) to +1 (positive).

### 3.4. Conditional logistic regression model of cumulative occupational exposures by agent family

Conditional logistic regression models identified significant associations for established covariates and nominal exposure associations (Figure 2A–F). In models combining PD and Parkinsonism versus controls, a parental history of PD or tremor was strongly associated with disease (OR = 4.55, 95% CI: 2.44–8.48; p < 0.001), and the SNCA rs356219 CC genotype was associated with increased odds of disease (OR = 2.17, 95% CI: 1.33–3.52; p = 0.002). Similar effect estimates for parental history were observed in PD-only models, whereas associations in Parkinsonism-only models were less precise due to fewer cases. The SNCA TC genotype was not associated with disease, and smoking showed an inverse association with PD versus controls (OR ≈ 0.58; p < 0.05). For cumulative occupational exposures derived from the ALOHA+-JEM (Figure 2D–F), high exposure to pesticides (vs unexposed) was positively associated with both combined PD + Parkinsonism (OR = 2.98, 95% CI 1.23–7.25; p = 0.016) and PD alone (OR = 3.56, 95% CI 1.25–10.15; p = 0.018). Odds ratios for metals, aromatic/chlorinated solvents, mineral dust, and gases and fumes were generally near unity with wide confidence intervals, providing no consistent evidence of association. None of the exposure categories remained significant after FDR adjustment (q > 0.05). All conditional logistic regression results, including FDR-corrected q-values, are reported in Supplementary Table S3.

**Figure 2.**
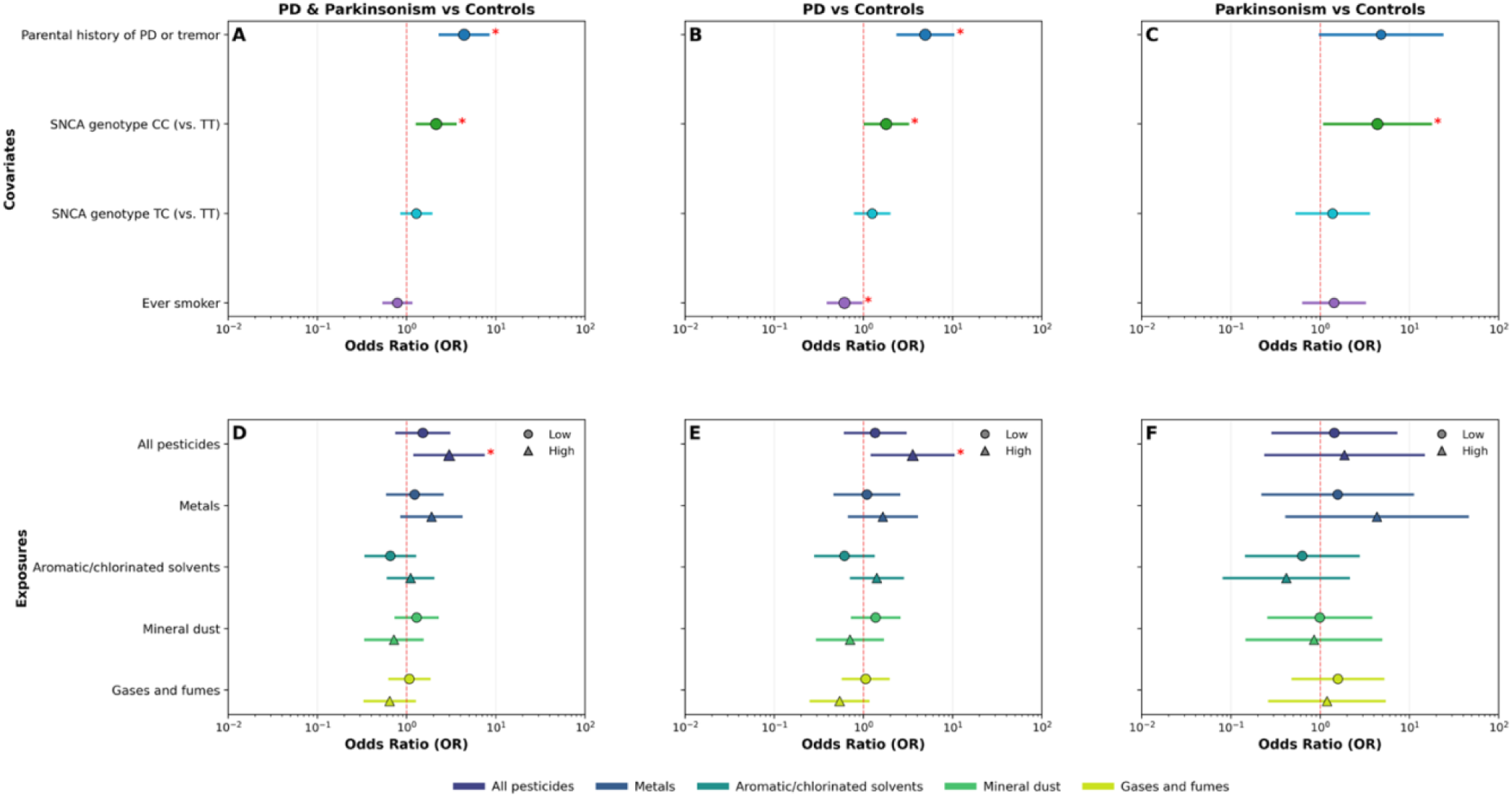
Conditional logistic regression for cumulative occupational exposures derived from the ALOHA+-Job Exposure Matrix (ALOHA+-JEM). Forest plots show odds ratios (ORs) and 95% confidence intervals for matched case–control analyses; the dashed vertical line marks OR = 1. Panels A–C display covariates included in the cumulative-exposure models, and panels D–F display cumulative occupational exposures derived from the ALOHA+-JEM. Results are shown for (A,D) combined Parkinson’s disease (PD) and Parkinsonism versus matched controls (N = 334 case–control pairs); (B,E) PD versus matched controls (N = 260 PD–control pairs); and (C,F) Parkinsonism versus matched controls (N = 74 Parkinsonism–control pairs). Covariates include parental history of PD or tremor, SNCA rs356219 genotypes (CC and TC vs TT), and ever smoker. Exposures include all pesticides, metals, aromatic/chlorinated solvents, mineral dust, and gases and fumes. Red asterisks (*) indicate statistical significance at p < 0.05 (uncorrected).

### 3.5. Single index weighted quantile sum (WQS) regression of cumulative occupational exposures

Single index weighted quantile sum (WQS) regression indicated a positive association between the cumulative occupational exposure burden index and disease status (Figure 3). For the combined PD and Parkinsonism model, the positive WQS index was associated with higher odds of case status (OR = 1.15; 95% CI: 1.00–1.30). The effect was similar in magnitude for PD alone (OR = 1.12; 95% CI: 0.90–1.33) and Parkinsonism alone (OR = 1.19; 95% CI: 0.86–1.59), although the wider intervals in the latter reflect reduced precision due to the smaller number of Parkinsonism cases. Statistical significance and stability were evaluated based on the variability of the repeated-holdout distributions, indicating the combined PD and Parkinsonism model as statistically significant. Component weights derived from 100 bootstraps and 100 repeated holdouts are shown in Figure 4. All pesticides received the highest mean weight in every WQS model, with mean values of 0.434 for combined PD + Parkinsonism, 0.400 for PD alone, and 0.277 for Parkinsonism. All three exceeded the equal-weight threshold (τ = 1/5 = 0.200) in their respective models, indicating that pesticides contributed more strongly than would be expected under equal weighting. Metals were the second-largest contributor, with mean weights of 0.210 (combined), 0.227 (PD), and 0.231 (Parkinsonism). Metals exceeded τ in the combined, PD, and Parkinsonism WQS models and variability across holdouts was moderate, reflecting a stable secondary contribution. The remaining components showed lower mean weights and remained below the equal-weight threshold. In the combined PD + Parkinsonism model, aromatic/chlorinated solvents had a mean weight of 0.185, mineral dust 0.100, and gases and fumes 0.071. In the PD model, aromatic/chlorinated solvents averaged 0.194, gases and fumes 0.099, and mineral dust 0.080. In the Parkinsonism model, mineral dust (0.188), gases and fumes (0.162), and aromatic/chlorinated solvents (0.156) had weights below τ. Overall, the cumulative occupational exposure burden index was positive across all WQS models, and the component-weight patterns consistently identified all pesticides and metals as the primary drivers of the exposure burden index effect. All WQS results are reported in Supplementary Table S4 and S5.

**Figure 3.**
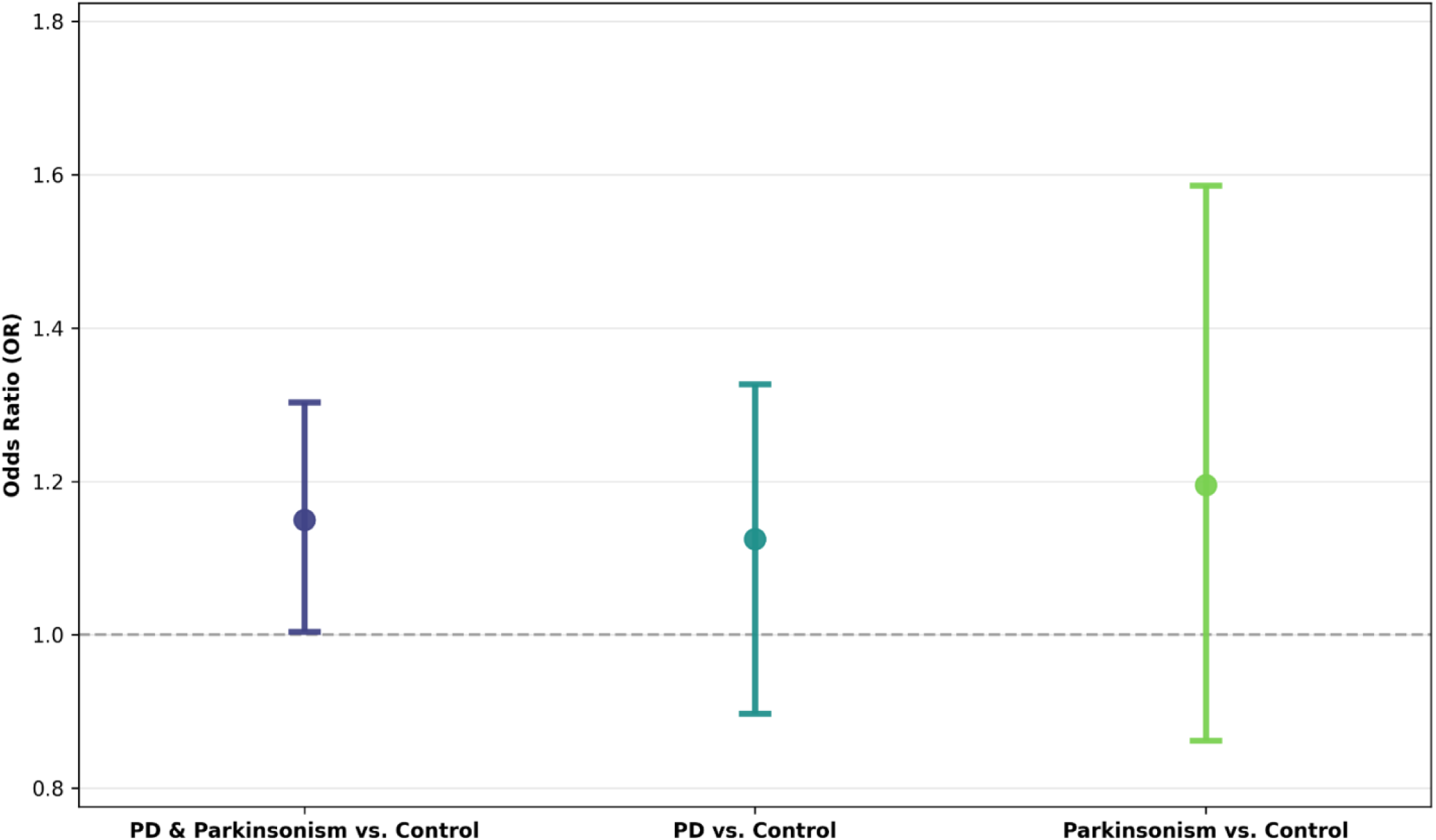
Weighted Quantile Sum (WQS) regression results for the cumulative occupational exposure burden index derived from the ALOHA+-Job Exposure Matrix (ALOHA+-JEM). Models were estimated using the gWQS R package with 100 bootstraps and 100 repeated holdouts, adjusting for ever smoker, parental history of PD or tremor, and SNCA rs356219 genotype. Bars represent odds ratios (OR) with 95 % confidence intervals for the positive WQS index. Analyses compare: (A) combined PD and Parkinsonism versus matched controls (*N* = 334 case–control pairs), (B) PD versus matched controls (*N* = 260 PD–control pairs), and (C) Parkinsonism versus matched controls (*N* = 74 Parkinsonism–control pairs). Confidence intervals were derived from the repeated-holdout distributions, and p-values were not computed.

**Figure 4.**
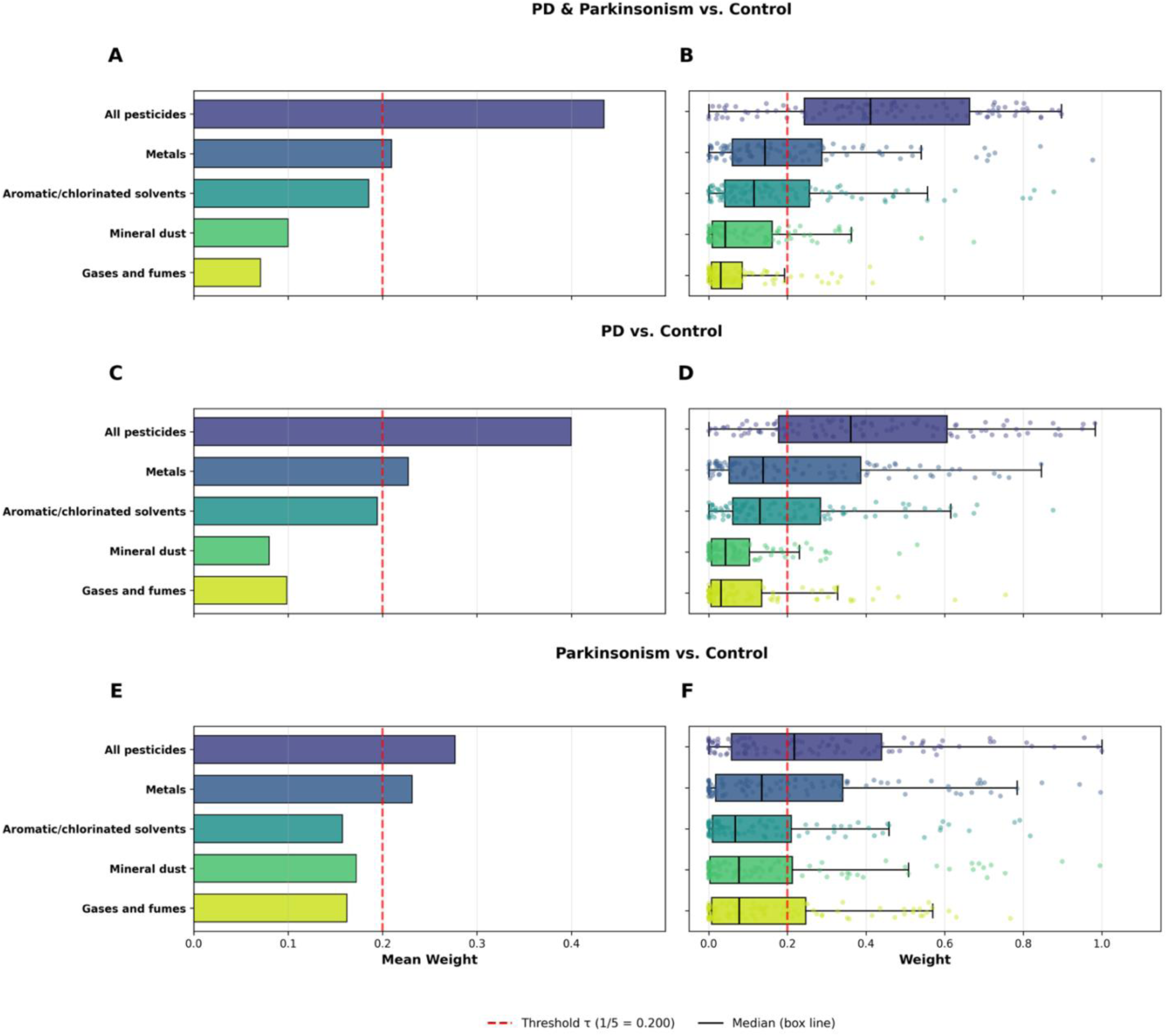
Weighted Quantile Sum (WQS) regression derived component weights for the cumulative occupational exposure burden index based on the ALOHA+–Job Exposure Matrix (ALOHA+-JEM). Models were estimated using the gWQS R package with 100 bootstraps and 100 repeated holdouts, adjusting for smoking status, parental history of Parkinson’s disease (PD) or tremor, and SNCA rs356219 genotype. All case groups were individually matched 1:1 to controls on age, sex, and occupational duration. Panels A, C, and E display the mean component weights (wᵢ) averaged across bootstraps for (A) combined PD and Parkinsonism versus matched controls (N = 334 case–control pairs), (C) PD versus matched controls (N = 260 PD–control pairs), and (E) Parkinsonism versus matched controls (N = 74 Parkinsonism–control pairs). The red dashed vertical line marks the equal-weight threshold (τ = 1/5 = 0.20), representing the expected contribution if all components contributed equally. Panels B, D, and F show the corresponding repeated-holdout weight distributions, where boxplots depict variability across holdout iterations. Points represent holdout-specific weights (N = 100).

## 4. Discussion

In this matched case–control study, we evaluated the association between cumulative occupational exposures and Parkinson’s disease (PD) and Parkinsonism using a job-exposure matrix–based approach. In conditional logistic regression models mutually adjusted for co-exposure, high cumulative pesticide exposure showed nominal positive associations with combined PD and Parkinsonism and with PD alone; however, these associations did not remain statistically significant after correction for multiple comparisons. Evidence for other exposure families, including metals, was less consistent. Additionally, In secondary analyses, weighted quantile sum (WQS) regression indicated a modest positive association between a composite occupational exposure burden index and combined PD and Parkinsonism, with pesticides contributing most strongly to the index and metals representing a secondary contributor. These findings should be interpreted cautiously, as WQS was applied to explore co-exposure structure and relative contributions rather than to estimate independent causal effects of individual exposures. Taken together, these results suggest that composite exposure-burden modeling may provide complementary insight to single-exposure analyses by incorporating information across multiple occupational exposure families.

The observed pattern for pesticides is consistent with a substantial body of toxicologic and epidemiologic evidence suggesting a role for pesticide exposure in Parkinsonian disorders. In a prior study applying the ALOHA+-JEM, van der Mark et al., did not observe statistically significant associations between pesticide categories and PD; however, elevated odds ratios were consistently observed in higher exposure categories for pesticides overall and for the functional subgroups, insecticides, herbicides, and fungicides. Analyses of selected active ingredients identified a significant association with the fungicide benomyl (van der Mark et al., 2014). Experimental studies further demonstrates that chronic, low-dose exposure to multiple pesticide classes can reproduce key features of Parkinsonian disorders, including dopaminergic neuron loss and motor dysfunction (de Souza Nascimento et al., 2025; Paul et al., 2024). Across pesticide classes, convergent neurotoxic mechanisms, such as oxidative stress, mitochondrial dysfunction, impaired proteostasis, and neuroinflammation, have been shown to disrupt dopaminergic pathways central to Parkinsonian pathology (Amaral et al., 2025). Together, this body of evidence provides biological plausibility for the nominal associations observed in the present study.

Although associations for metals were not consistently observed in the primary regression analyses, the contribution of metals to the composite exposure index in WQS models suggests a potential role for cumulative metal exposure in the broader occupational exposure context. This interpretation is consistent with a substantial body of mechanistic and epidemiologic evidence linking metal exposures to Parkinsonian outcomes, particularly for phenotypes extending beyond idiopathic PD. Evidence from occupational cohorts, including welding and mining populations, indicates that chronic manganese exposure is associated with parkinsonism-like motor symptoms that often manifest at younger ages (Khindri and Maj, 2025). More broadly, occupational settings frequently involve cumulative metal burdens accrued over work histories, with studies implicating manganese, lead (including cumulative bone lead), iron dysregulation, and other metals in nigrostriatal vulnerability (Lucchini and Tieu, 2023; Pyatha et al., 2022; Zhao et al.,2023). Mechanistically, metals have been shown to disrupt neuronal integrity through oxidative stress, mitochondrial dysfunction, dysregulation of metal homeostasis, and promotion of α-synuclein misfolding and neuroinflammation (Chen et al., 2025; Shvachiy et al., 2023). While these pathways provide biological plausibility, the lack of consistent associations in single-exposure models in the present study suggests that any contribution of metals may be modest or more readily detectable when considered as part of a broader cumulative exposure profile.

This study has several important strengths. It is a matched case–control investigation of PD and Parkinsonism using a harmonized, expert-reviewed JEM (ALOHA+-JEM) to assess occupational exposures across multiple agent families. The use of detailed lifetime occupational histories enabled estimation of cumulative exposure across the working life, providing a more comprehensive representation of long-term occupational exposure patterns than approaches based on single jobs or cross-sectional exposure measures. All occupations were systematically coded using ISCO classifications and linked to the ALOHA+-JEM, allowing standardized exposure assignment across participants. While occupational histories were self-reported and may be subject to recall error, exposure assessment was conducted independently of disease status using a predefined JEM, reducing the likelihood of differential misclassification between cases and controls. Additional strengths include clinically confirmed diagnoses by movement-disorder specialists, harmonized evaluation across specialized centers, and matching on age, sex, and occupational duration, which improves comparability between cases and controls and helps control for these factors by design. Finally, the use of WQS regression provided a complementary, exploratory approach to characterize co-exposure structure and relative contributions of multiple occupational exposures.

Several limitations must be acknowledged. First, exposure assessment was based on a JEM, which assigns exposure at the group level and cannot capture variability in exposure intensity across specific tasks, workplaces, or time periods within the same job. As a result, exposure misclassification is likely and may bias effect estimates, although the direction and magnitude of such bias cannot be directly assessed in the present study. Second, the hospital-based case– control design introduces potential for selection bias, as controls may not fully represent the source population from which cases arose. In addition, differential participation or response rates between cases and controls could affect comparability, and occupational histories were self-reported, raising the possibility of recall error. Although exposure assignment was conducted independently of disease status using a standardized JEM, reducing the likelihood of differential misclassification, the validity of exposure information remains a limitation. Third, residual confounding cannot be excluded. Although models were adjusted for key covariates, unmeasured occupational and non-occupational factors, such as rural residence or environmental exposures correlated with pesticide use, may influence observed associations. In addition, reciprocal confounding among correlated occupational exposures may persist despite mutual adjustment in regression models and complementary WQS analyses. Fourth, the definition of exposure based on broad agent families lacks specificity and may obscure heterogeneity in risk across individual agents. This limitation is particularly relevant for pesticide exposure, which is often intermittent and variable over time compared with more continuous exposures such as metals or dust, potentially increasing susceptibility to misclassification and confounding. Finally, certain toxicologically relevant agents, including specific solvents and individual pesticide active ingredients (e.g., paraquat), could not be evaluated. Future studies incorporating biomonitoring, agent-specific exposure assessment, and longitudinal designs will be important to better characterize exposure–response relationships and underlying mechanisms. Overall, the findings of this study should be interpreted cautiously. While results are consistent with a potential role for occupational exposures in Parkinsonian outcomes, the semi-quantitative nature of exposure assessment and the observational design limit causal inference.

## Data Availability

All data produced in the present study are available upon reasonable request to the authors.

